# Gender-based social norms, psychosocial variation, and power relations during delivery and post natal care: a case study qualitative research in rural Ethiopia

**DOI:** 10.1101/2022.12.25.22283934

**Authors:** Ketema Ayele, Yohannes Kebede, Lakew Abebe, Abebe Mamo, Morankar Sudakar

## Abstract

**Background:** The World Health Organization (WHO) strongly encouraged men to support women to use maternal health care services. However, especially in developing countries, maternal health care has traditionally been viewed as a women’s issue, men making little or no contribution, despite the fact that sexuality and children are shared products. The study’s goal was to understand how gender-based norms, psychosocial variation, power relations, and community social support are related to child delivery and post-natal care services.

**Method:** The study took place in three rural districts of Ethiopia’s Oromia regional state, Jimma Zone. An in-depth interview and focus group discussion were held with a group of carefully chosen health professionals, community health development armies, and religious leaders. The data was collected, translated, and transcribed by men and women with experience in qualitative research. For data analysis, Atlas ti version 9 was used. Gender-based delivery and postnatal care service utilization were investigated using gender-based categories.

**Result:** Independent and shared gender-based roles were identified as a means to improve maternal health-care services delivery. Men have the ability to persuade pregnant women to use delivery services and post-natal care. The place of delivery is determined by the levels of gender power relations at the household level, but women are usually the last decision-makers. Furthermore, the community’s belief that giving birth in a health facility makes women look clean and neat, as opposed to home delivery, increases their intention to use maternal health care services..

**Conclusion:** Understanding men’s and women’s roles, knowledge levels, beliefs, decision-making, and social support significantly improves maternal health care service utilization, allowing pregnant women to receive delivery services at a health facility. Pregnant women were the last to make a decision about using maternal health care services. There are still gaps in knowledge and negative beliefs about maternal health care that limit delivery and postnatal service utilization.

**Recommendation:** Increasing community-level social support can significantly improve maternal health care service delivery. Men’s concern about institutional delivery is increased when they are viewed as the primary audience during maternal health education. Researchers should focus on the mechanisms through which men participate in delivery in the future, and postnatal services will fully address men’s involvement in maternal health care services.

## INTRODUCTION

Gender is defined as what society believes about people’s appropriate roles, duties, rights, and responsibilities, as well as their attitude, value, relative power, accepted behaviors, and opportunities based on their sex **(1)**. It is one of the common social determinants of maternal health that includes various ideas in it, basically about how women and men interact and the nature of their relationships **(2)**.

Various strategies and initiatives support the issue of gender in maternal health care; the impetus for the initiative was the 1994 International Conference on Population and Development (ICPD) held in Cairo, Egypt, which mentioned that males should be involved in various maternal health service utilizations (3-6). They also supported the (1995 conference on World Women’s Day in Beijing, China (7). In the Addis Abeba Declaration, the concept of men’s involvement in maternal health was one of the issues **(8). T**he World Health Organization (WHO) in 2015 strongly recommended and declared there was low quality evidence of male involvement in maternal health care service use. They mentioned men should be involved in maternal and child health-care services **(9)**. Although the global MCH declarations and initiatives consider gender-related roles to have a significant influence on maternal health care, limitations have been widely observed in the mechanisms of gender integration in maternal health care.

Despite a 44% reduction in maternal mortality rates globally, targets for 2015 were not met, particularly in low- and middle-income countries **(10)**. Maternal mortality, for example, was 19 times higher than in developed countries **(11)**. In 2015, globally, 303,000 women died of preventable causes before, during, and after giving birth, which is 1 in 180 women **(12)**.

**I**n Ethiopia, according to Ethiopia Demographic Health Survey (EDHS) information, the maternal mortality ratio in 2019 was 224 per 100,000 live births **(13)**. It is continuing to become one of the major public health issues. Gender inequity in maternal health is not fully addressed when only dealing with women. Unless men and women are considered part of the causes and the solution, it is hard to successfully challenge and transform gender norms to improve the desired outcome of maternal health care services, but understanding the contribution of gender to integrate in maternal health care will help to ease the burden on women during maternal health care **(14)**. A study in Nepal concludes that greater male involvement has the potential to improve maternal and child health conditions**(15)**. Therefore, global evidence, declarations, and national strategies support the issue of gender in maternal and child health care services. So, asking the question, “Why is gender equity in maternal health care during delivery and post-natal care not ensured as expected?” would have been critical to ensuring entrenched challenges in maternal health care services. The research findings in Afar and Kefa, Ethiopia, revealed the role of gender: men are dominant decision-makers for maternal health care and did not allow her to attend meetings with the health development army due to the fear that she could have known her rights **(16)**.

As the literature shows, gender-based power relations have a significant impact on access to delivery and postnatal services **(17)**. However, emerging evidence and program experience indicate that engaging men during delivery and postnatal care could have considerable health benefits for women in low- and middle-income countries **(18)**. A study conducted in Uganda identified husbands who didn’t notice the involvement of men as a key predicament in maternal service utilization. Whereas women reported that their husband’s lack of involvement hampered their use of delivery and postnatal care services **(19)**. In Malawi, the process of childbirth is an issue for women only; the husband should not know about childbirth **(20)**. In Ethiopia, various stakeholders in maternal and child health from zones to the federal level identified that men have a role in doing physically intensive tasks, while women are more engaged in care giving, household tasks, and health promotion tasks. Women also had low health seeking practices because men tended to act as gatekeepers for maternal health service use by women **(21)**. Many of the studies didn’t consider the internalized beliefs and subjective experiences of men, which is called a transformative approach, rather than focusing on the observable behavior of men, which is taken as men’s involvement in fulfilling gender equity to improve maternal and child health outcomes **(22)**. Besides the observable roles of men, such as physical support and gender relations, the subjective experiences of men were not considered a strategy in reductionist and instrumentalist approaches **(23)**. Therefore, to manage entrenched gender-sensitive maternal health care, we must consider both men’s and women’s subjective experiences with maternal health care.

According to the recommendation of the World Health Organization, male involvement during child birth and after birth contributes a lot to facilitating support, improving self-care, and improving maternal and child health outcomes. Because men are considered the gate-keepers and decision-makers for prompt access to MNH services both at household and community level, no matter how the importance of male involvement in the continuum of maternal health was acknowledged, there was no quality of evidence produced in low and middle-income countries from the community and health professional perspectives **(9)**.

Without considering the reason behind, male attendance during delivery and saving money for transportation during an emergency are considered men’s involvement. However, to clearly describe male involvement understanding the motivational and deriving forces of men has enormous contribution to address gender inequity **(24)**.

A study in Nepal found that the husband’s role is complex and multidisciplinary, including but not limited intervening to assist their wives in maternal health, including giving birth in the health institutes. Some also acknowledge reducing work load, particularly in harsh environments **(15)**. The gender role at household and community levels improves the intention of women to utilize delivery services **(25)**. The desire of men to be involved was improved and described in association with social attitudinal change **(15)**. The qualitative study of African American parents in the USA mentioned that there is equal investment and interest in having a child, and that the required responsibility of having the child is willingly shared between the two parents **(26)**. On the other hand, male involvement, on the other hand, would be reduced if they were perceived as a rule, descended from the government, which also stigmatized women who did not have a husband **(20)**. Another study revealed that men are commonly involved in the first ANC visit and also have a responsibility to give the women birth in the health institute **(20)**. According to an Indonesian study **(27)**. Men should have the knowledge to improve maternal health care utilization. In Uganda, various stakeholders engaged in maternal health agreed on the importance of male involvement in maternal health **(28)**. However, in some cultures, men’s involvement in maternal health care is considered by the peer group as a weakness. It has also been perceived as a loss of women’s rights to make decisions by themselves **(29)**. similarly participants in another study perceived as a means to express their love for the pregnant woman **(20). As** maternal health care stakeholders expressed, it was men who decided whether women should use maternal health care **(21)**. Whereas in Nigeria, men have no interest in attending the delivery service but provide financial and material support. However, the World Health Organization recommendation considered that men’s involvement should be promoted in maternal and child health care, provided that they respect, promote, and facilitate women’s choice and their autonomy in decision-making **(30)**.

The aim of this study is to explore gender-based roles, psychosocial variation, gender power relations, and social support during institutional delivery and post-natal care from community and health professional perspectives in Africa, particularly Sub-Saharan Africa (SSA) **(15)**. As a result, it is critical to investigate in rural areas of developing countries such as Ethiopia, where high maternal mortality ratios will provide guidance for policy and program development that will aid in the integration of men into maternal health care service utilization.

The findings presented in this research were a component of the Innovative Maternal and Child Health in Africa (IMCHA) project, which was implemented in Jimma Zone, Ethiopia. The overall objective of the project was to achieve better maternal and child health through applying information, education, and communication as well as improving the maternal waiting areas. In one of Ethiopia’s nations, male dominance is widely observed. In contrast to other reproductive health issues where male involvement significantly contributes to service utilization and maternal health improvement, there was little or no evidence of male involvement in the continuum of maternal health service utilization evidence available despite it being recommended by WHO and Ethiopia’s national maternal health policy.

## 2. Methods and materials

### 2.1. Setting

The study was conducted in three districts of Jimma Zone, which is located in Ethiopia’s south-west region. The rural districts are very far from the city of Jimma Zone, with limited infrastructure in roads, communication, transportation, and health facilities. The health facilities in all selected districts are not equally accessible. In all selected districts, the total populations were estimated at 180,000 to 270,000 in 2016, according to the central statics authority of Ethiopia. The predominant ethnic group is Oromo, and the religion is Muslim.

### 2.2. Study design

This study used a qualitative case study design approach, which is appropriate to explore community including husband and wife and health professionals’ perceptions of gender-based norms, psychosocial variations, power relations, and social support about delivery and post-natal care services.

### 2.3. Participants and sampling

Purposive sampling was used to select study participants, both men and women, based on their role in the community. They have a capacity to understand the community’s gender-based norms, levels of psychosocial variations, power relations, and social support about delivery and post-natal care services. Besides this, they were selected based on the role they are playing in the community, such as providing health support, counseling, and acting as a leader in the community. The participants were the women’s health development army (WDA), the male health development army (MDA), religious leaders (RL), health extension professionals (HEP), midwifery nurses, and primary health care unit directors (PHCUD).

### 2.4. Data sources

The data was obtained from the IMCHA project and collected from three rural districts in the Jimma zone, such as Gomma, Seka Chekorsa, and Kersa.

### 2.5. Data collection methods

Focus group discussion (FGD) was held with the Women’s Development Army and the Men’s Development Army and in-depth interviews carried out with religious leaders, health extension professionals, primary health care unit directors, and midwifery nurses. A semi-structured guide, an in-depth interview, and focus group discussion were employed to explore the data.

Basically, the study was conducted by Innovative Maternal and Child Health in Africa (IMCHA) as part of the Safe Motherhood Project, with the aim of improving maternal and newborn health by scaling up interventions in Africa. The joint project was led by Jimma University and Ottawa University in Canada. A five-year project started in 2016 and will end in 2022. The data collection process was held using skilled male and female qualitative data collectors, who had been recruited and took a five-day theoretical and field practice training. The interview and discussion guides were checked to ensure they were compatible with the local socio-cultural context. A pre-test was conducted other than the actual study areas, and an indispensable modification was made. At a convenient time and location, data were collected separately from male and female participants. The project’s goal was to improve maternal and child health; however, qualitative data was gathered from community groups and health professionals who were concerned about maternal health care services. A total of 32 in-depth interviews and focus group discussions from three districts and six community groups took place. For this particular research, 12 focus group discussions (FGD) from three districts and 20 in-depth interviews from three districts were used. FGD were conducted with the male health development army (MDA) and women’s health development army (WDA), as well as in-depth interviews with religious leaders, health extension workers, midwifery nurses, and primary health care unit directors (PHCUD). Saturation of data was the reason for limiting the number of interviews and discussions with the participants. Both interviews and discussions were held for an average of 75 to 120 minutes, respectively, with an average of 90 minutes. Respondents were selected purposefully from each community group except health extension professionals, midwives, and PHCU directors due to their limited numbers in the districts. The safe motherhood project has aimed to roll out two interventions (an information, education, and communication intervention and MWA improvement) and has conducted multiple rounds of data collection as a baseline prior to intervention and an end line after post-intervention. This study focused on the baseline data to compare with the endline data after the intervention of the IEC on maternal health care services and the upgrading of the maternal waiting area with essential equipment.

### 2.6. Data analysis

Male and female data collectors translated and transcribed the collected data immediately after data collection. The transcribed data were analyzed using Atlas ti ver 9, qualitative analysis, and statistical software. The entire FGD and key informant interviews were exported to the software, which created a new project. Initially, the data was coded by two coders based on the participant’s perceived gender-based descriptions. Then codes with similar ideas merged together and formed categories such as gender-based roles, knowledge, belief, social support, and decision-making on maternal health care services. The coding strategies used to answer the research question were descriptive methods because the analysis was performed to explore the descriptions of the different health professionals and segments of the community about the delivery and post-natal health care services used. However, in their descriptions, they mentioned the social norm, the psychosocial aspects, and power relations.

### 2.7. Trustworthiness

To ensure the rigor of this research, an ample number of criteria were critically considered. To begin with, adequate data from various community and health professionals was collected to produce significant findings. Second, adequate time was spent in the field to obtain trustworthy results. Third, we identified theoretical goals that should alienate the context of the study subjects. Fourth, I followed a strict procedure when conducting interviews, writing field notes, and analyzing data. The four basic qualitative research trustworthiness measures were considered during data collection and analysis. The first is credibility; to achieve this, data were collected by experienced teams of data collectors; data saturation was considered; data were transcribed and translated on time; and there was extensive field engagement. Additionally, thick descriptions were provided for the categories of gender perspective and supportive quotations, which added value to the content. Aside from that, multiple data collection methods from various groups of participants (multivocality) were used to construct a multi-faceted, more complicated, and more credible picture of the result; the second is dependability, to ensure peer debriefing and audit trials were performed during data collection, translation, and transcription; and the third is transferability, because the data were collected in a resource-limited setting and multiple data-data triangulation was used; and the fourth is transferability, because the data were collected in The findings of the research have a methodological and practical contribution to the body of knowledge of gender-based maternal health care since there is limited evidence in this field of study; the fourth is conformability, to ensure the process of data analysis (interpretation) is strictly grounded in the data to avoid the preferences and view points of the researcher.

### 2.8. Ethical considerations

Potential respondents were given verbal information about the aim of the study and invited to participate with their willing consent. Consent was obtained verbally by the data collectors just before the time of interviews. Filed notes and recordings on tape were made to remember the natural setting and proper transcription. To ensure confidentiality and anonymity, the participants’ names were not mentioned; rather, their age, number of children, educational status, and duration of service in each particular social and professional group were mentioned. The FGD was undertaken at the participant’s residence at a convenient time before they started their daily activities, whereas the in-depth interview was conducted at the participant’s offices. A limited amount of money reimbursed for transportation purposes was provided. Ethical approval was obtained from Jimma University Research Ethics Institutional Review Boards (JUEIRB) and the University of Ottawa.

## 3. Results

### 3.1. Participant profiles (age, religion, category data source, and male, female)

A total of 95 female and 97 male participants from the community were involved in the focus group discussion, whereas in the in-depth interview, four religious leaders, four HEWs, four PHCUDs, and four midwifery nurses were involved. The mean age of male community participants was 41 years old, and 35 years for women. The educational status of the participants ranges from no formal education to a university degree. The number of participants’ children ranges from no children to 10 children.

### 3.2. Gender-based category development

The following gender-based roles, psychosocial variations, and power relations categories about delivery and post-natal care services were explored systematically following the procedure during the analysis. Then the transcribed in-depth interview and FGD data were exported into Atlas ti Ver 9, for a second fast reading of the entire document to determine the codes. The codes had been given based on the participant’s gender-based explanation that fit into the predetermined categories. Then, after related codes merged together and formed sub-categories, at last, very similar sub-categories merged together and produced five categories.

### 3.3. Description of gender-based categories during delivery and post natal care

In this qualitative study, a total of three well-defined thematic categories were identified: 1. Gender-based norms during delivery and the post-delivery period; here, we basically addressed men’s and women’s roles that were carried out to facilitate and prevent the utilization of maternal health-care services. 2. Gender psychosocial variations, including gender-based knowledge; under this category, variation and similarities of gender-based knowledge during delivery and postnatal care services, as well as gender-based belief; under this category, gender related beliefs were either supportive or hindering of the utilization of delivery and postnatal services. 3. Gender-based power relations: under this category, unindependent and collective decision-making that facilitates and hinders the delivery and postnatal care services were included.

### 3.4. Gender-based norms during delivery and PNC

Gender-based maternal health care norms embrace an array of gender pertaining to delivery and postpartum care as an individual and collective role based on community and health workers’ perspectives. Commonly, men and women are concerned about the better health outcomes of pregnant women and newborns.

#### 3.4.1. Gender norms identified to women during labor and delivery

The role of the pregnant woman as an individual during labor and delivery has been mentioned in two paradoxical ways based on maternal health-seeking behavior. The first is that women who had an intense interest in giving birth in the health institutes went there before the true sign of labor had started, in order to get proper follow-up and adequate rest. On the other hand, pregnant women were classified as those who remained at home till the true sign of labor had started; if possible, they preferred to give birth at home; otherwise, depending on the condition, they would go to the health facility.

> *There are women who come at the initiation of their false labor and stay here [at the health facility] for one or two weeks up to their delivery time…*…*the others come when labor has failed at home*. ***[PHCUD, Kersa District]***

> *They [pregnant women] come to the health center just when the labor starts; before that, they prefer to give birth at home. So, giving birth at a health center is considered the second option*. ***[Husband, Seka district]***

> *There are pregnant women who are refusing birth at the health center*. ***[Religious leader, Seka District]***

> *Those pregnant women who decided to give birth at home do not tell Shane [one to five women network] members. Rather, they simply keep quitting, and the entire family says that she [the pregnant women] is fine*. ***[WDA, Gomma District]***

Women’s collective gender norms that facilitate institutional delivery encompass many activities, including but not limited to escorting pregnant women to health institutes, doing household chores, and providing proper care for the kids who were left at home when the pregnant women went to the health facility for delivery.

> *Some women have gone with her [laboring women]; they prepared a coffee ceremony and drank it while she was in health care. She has been morally supported until her return to her home. She didn’t suffer from anything while she was at the health center. [Husband, Gomma district]*

> *Those women who left at house prepared the place where we takes rest after returning home, as well as anything else at home; for example, preparing coffee, cooking food, and making all things at home ready. [Women who gave birth, Kersa district]*

> *The other women who are still at her home clean the house as well as stay with her children and manage all the necessary things at home. [WDA, Seka district]*

#### 3.4.2. Gender norms identified to men during labor and delivery

Men’s role (including husbands) basically focuses on facilitating transportation to carry the laboring women to the health facility, buying food and medicine, and escorting them to the health facility during delivery. The intention that the husband advises and supports his wife to give birth in the health institutes was classified into two ways; the first is to save the life of the woman and the newborn. The second is, to refrain from taking family responsibility alone in case the pregnant women die during delivery at home.

> *The husband advises her [pregnant wife]… because he is afraid of the problems and expenses that may come with childbirth. The husband has the feeling whatever maternal problems happen; it will affect or hurts his family first. Therefore, he needs to take care by taking the pregnant women to health facilities. [Husband, Gomma district]*

Besides these, men at the household level are responsible for informing [communicative role] the community leader early when the labor sign has started, they could mobilize the community members in case of no ambulance service to carry laboring women to health facilities or to the main road in the traditional way.

> *“What is expected from the husband is only asking for assistance [the community], then they inform each other and easily come for help*.*[RL, Gomma and Seka districts]*

> *The husband tells to EDIR (local traditional support system using regular money collection) leader, so the leader will mobilize the public. Then the people immediately carry the pregnant woman to health care using traditional hand-crafted beds. [MDA, Gomma District]*

> *…. we all [male community members] are with her once the labor begins. We go with her till her arrival at the health center. We attentively stay there to follow her condition and whether she is being referred or not. We never go to our daily work that day. Every member of MDA, the neighbor, and the community at large should attend her. We do whatever is needed; we carry if there is no ambulance. [Husband, Gomma District]*

#### 3.4.3. Gender norms identified to women during PNC

There were very few women-specific roles identified during postnatal time, with the focus rather on child care such as breastfeeding, which is the reproductive responsibility of the women for the new-born baby. This was due to the cultural restriction of women moving outside of the home after delivery, and because most women and the new-born baby didn’t experience health problems. Most of the activities were performed by the family members, including the husband and the neighbor. However, it was possible to see in three categories how the women perceived postnatal care attendance: the first was women who remained at home until they reached 40–45 days without receiving the postnatal care most commonly associated with the distance from the health facilities. The second category of women were not visited the health facility but HEW visits them despite it was not considered as PNC. The third group was, even if their number were very few who dwell near to the health facilities have visited at the seventh day of post delivery.

> *The reason the women do not use PNC is because they think themselves and their baby are healthy and decided to come for immunization only. [Midwifery, Gomma District]*

> *“Women who understand the importance of the PNC service could use it. [PHCUD, Seka District]*

> *The mother and the baby will be free of all the problems, as we [HEW] have seen during PNC. [HEW, Seka District]*

> *There is a traditional belief that obligated women who gave birth not to be outside for 40–45 days after delivery. So, we [midwifery] recommend health extension workers conduct PNC-2 visits. [Midwifery, Gomma District]*

#### 3.4.4. Gender norms identified for men during PNC

The men’s specific role during post-delivery time was mostly associated with fulfilling all the necessary food stuffs for the women who gave birth and the family as a whole, which was the continuation of pre-delivery preparation. In general, the husband’s role is to fill all the gaps that existed during the postnatal period to maintain the dignity of the family.

> *He [the husband] is also concerned about the food she will use after delivery and the clothes that she wears. [WDA, Gomma District]*

> …*so that the husbands make ready and clean the bedroom where she will stay (after delivery). [Husband, Seka District]*

The social norms urged men to provide proper meals, dresses, and sanitation to the women who gave birth, unless the community will humiliate them in different ways, like insulting them. Therefore, men were involved in the household chores when another individual provided hospitality services for the guests who visit and congratulates the newborn woman.

> *He (the husband) has a fear that friends and neighbors will laugh at him for the lack of things at home like clothing as well as anything else while visiting them after his wife gives birth. Therefore, he supports his wife very much. [Women who gave birth, Seka District]*

> …*if they [parents] fail to get a person who supports her in doing things while she is at rest (post delivery), the husband is the one who does that*.*[WDA, Gomma District]*

### 3.5. Gender-based power relation during delivery

As most participants have mentioned, gender-based power relations have been classified into three major categories: first, the shared decision-making of the parents, including other very close relatives. Second, the decision was made by the pregnant woman alone, and despite the parents’ common discussion, her decision was approved at last to select the place of delivery. Third, the decision was made by the men alone. The men have made decisions either on behalf of the women when they were found in stressful situations during delivery time or based on their own interests. However, there were socially disagreeable and controversial issues about the last decision-maker to select the place of delivery.

#### 3.5.1. Shared gender-based power relations to select the place of delivery

Commonly, both men and women partners held discussions and reached shared agreements about the place of delivery, often times in a health institution. Besides the shared decision-making roles of the parents, sometimes families of both parents and health extension professionals were involved in the process of decision-making, particularly to give birth in the health institutes. However, since husband and wife are living together initially, they held discussions and reached consensus on the place of delivery before moving on to the next steps of the decision-making process.

> *For deciding the place of delivery, she [the pregnant woman] talks with her husband and family, so that she can do as she prefers. Even so, he [pregnant woman’s husband] is the one who let her go to the right health facility as soon as she felt pain at this time. Therefore, husbands are not denying women the right to go to a health facility while they face labor; rather, they take her to the facility. [Women who gave birth, Seka District]*

> *Her [the pregnant woman’s] parents, his [husband] parents, and HEWs also participate in the decision to encourage the pregnant woman to go to a health center [at the time of labor]. [Husband, Kersa District]*

> *Even if she has the ability to decide where to give birth, the neighbor also decides for the pregnant woman*… … *the husband is also behind her and protects her from any harm. [WDA, Gomma District]*

Despite the fact that this was a very rare case, men didn’t accept the decision to give birth in a health facility. Therefore, women could move on to the next steps, either informing their neighbor or the one-to-five network group, in order to receive assistance in getting to the health facility.

> *Generally, she [pregnant women] decides with her husband; if he denies her the right to give birth at a health facility, she again decides with her neighbor and asks them to take her to a health facility. [WDA, Seka District]*

#### 3.5.2. Women decision making power during delivery service

As women discussants and religious leaders described, if the discussion between men and women partners failed and didn’t reach agreement, pregnant women’s would have been the last decision-making to select place of delivery. They rationalize her decision in terms of “*after all, it was her life that might be in danger; therefore there was nothing that impeded the woman from making the decision of institutional delivery*.”

> *The pregnant woman herself has to make the final decision. Currently, pregnant women are starting to make the decision by themselves. After all, it is about her health and life. [RL, Seka District]*

> *Pregnant women by themselves make the decision [for institutional delivery]. The husbands accept what the pregnant woman says*… *……… At this time, they have been using the health center for their own interests. [RL, Gomma District]*

> *As she preferred to give birth at a health facility, nothing hinders her from doing so since it is her right. Regarding her decision, she starts talking and discussing with her husband by saying, “I want to go [to] a health facility for delivery,” so she says to him [husband] “Either you take me to a health facility or I will go by myself” while she faces labor. Then, if he said no, she [the pregnant woman] would need to go to the health facilities; she would go by herself and give birth there at the health facility. [WDA, Seka District]*

#### 3.5.3. Men decision making power during delivery service

Even if the cases were very few, still men [husbands] have made decisions alone in the form of motivation and encouragement of the pregnant women about the place of delivery. There were tendencies that the women respected his decision. On the other hand, since the women are in a stressful and tension-filled situation at the time of labor, men could have made the decision on her behalf to carry her into the health institutions without requesting her consent. And in other situations, he denied the preference of the pregnant woman’s health institution for delivery.

> *The one who makes decisions is the husband. The woman herself can also decide, but the final decision will be made by the husband, who gets respect and is implemented. However, the husband’s decision is just to motivate and encourage her to attend the health center. [RL, Kersa District]*

> *There are husbands who bring their wives to the maternity waiting area without the consent of the women. [Midwifery, Gomma District]*

> *Sometimes mothers [pregnant women] cannot do anything alone. For instance, if she requests that her husband bring her to the health center, he might not bring her. [PHCUD, Seka District]*

> *There are still some pregnant women who say, “That is the decision of my husband, and if he says that you [the women] have to give birth here at home and don’t go to a health facility, she will give birth there at home by his will. Therefore, she is obeying what her husband ordered and not her own interests*. ***[Women participants, Gomma and Seka districts]***

### 3.6 Gender-based psychosocial variations about delivery and postnatal care

The gender-based psychosocial component comprises men’s and women’s knowledge and beliefs about delivery and postnatal care services. The knowledge and beliefs of both men and women have shown positive change, whereas still some knowledge gaps and negative beliefs were observed about maternal health care services.

#### 3.6.1. Knowledge of women identified during labor and delivery

Previously, women experienced morbidity and mortality due to a lack of awareness about maternal health services and the accessibility of those services. However, as participants mentioned, currently the vast majority of women’s have a better understanding of the benefits of giving birth in health institutions and the untoward effects of home delivery. Moreover, the two most common reasons they mentioned were fear of bleeding and saving the lives of women and newborns.

> *“*..*They (pregnant women) say that “there is no bleeding if we give birth in a health center*.*”* [PHCUD, Kersa District]
>
> *Mostly, they prefer to give birth at a health center. Due to the risk of giving birth at home and the safety and care provided by professionals at a health center, they prefer a health center*. [MDA, Kersa District]

They (pregnant women) know the risks related to delivering at home and the benefits of delivering at health facilities. Midwifery, Seka District]

> *“…the reason is,that, first, there is excessive bleeding during delivery. So; in order to prevent this; they prefer health care*. [MDA, Gomma District]

> *“The reason why they [pregnant women] are not giving birth at home is; they are getting advice*.*” “So, the concern for their health makes them attend health care during delivery*.*”* [MDA, Gomma district]

Particularly according to the perception of health professionals, the vast majority of pregnant women have a clear knowledge about the difference between giving birth at home and in health institutes; therefore, they have an improved intention to attend the maternal waiting home, even before the true labor sign has started; sometimes without waiting for anyone else’s assistance, even her husband can find her in the health institute’s maternal waiting area. Most of them were at the health facility at the time of delivery; they would not have been at home.

> *There are pregnant women who come at the time of the initiation of their false labor and stay here [health facilities] for one or two weeks up to their delivery time. This is due to the awareness they have…*.[PHCUD, Seka District]

> *After knowing the benefits of health services, pregnant women run to a health facility for delivery as soon as they face labor without waiting for anyone to take them. [WDA, Kersa District]*
>
> *They [pregnant women] know the risks related to delivering at home and the benefits of delivering at health facilities. …. [Midwifery, Seka District]*

The community perceived home delivery now a day associated with a sign of backward culture. There were absolute knowledge improvements about the importance of giving birth in the heath institute.

> *Currently, giving birth at home is considered* ***traditional or backward culture***. *When she gives birth at home, there are many challenges, like the overflow of blood, and the deaths of the child and mother. Delivering at health center saves her life and the newborn, gets good care, and the delivery system is safe*. [MDA, Kersa, District]

The idea was supported as **men felt ashamed when pregnant wives gave birth at home. The community members could insult him due to his irresponsibility** in taking care of the pregnant women and his failure to provide advice about giving birth in the health institutes, in case they knew the women had given birth at home.

> … *[Community] ask her [women] where she delivered [gave birth]; if she delivered at home, we [the community members] insult the husband; why he did this? All in all, giving birth at home is considered a backward culture, and people are becoming familiar with giving birth at a health center*. [MDA, Seka District]

Similarly, as participants expressed, the knowledge of women about institutional delivery was associated with the ways of evaluating health facility delivery in terms of the cleanliness **and neatness** of the women after completing the delivery services, with minimal blood spillage unlike home delivery. Another discussant agreed, saying that it was only a knowledge deficit that prevented the women from giving birth in the health institutes.

> *They are giving birth at a health facility by considering no loss of blood at delivery and delivering neatly, unlike previous times where there was high bleeding up to the extent that it spoiled the odor of the women*. [WDA, Kersa District]

> *They [pregnant women] prefer to give birth here at the health post, as they don’t have anywhere else to go for delivery. The reason for preference is to save one life from death; [WDA, Kersa District]*
>
> *Except a lack of knowledge, nothing hinders women from using health services, as they can all access them. [WDA, Seka District]*

> *After knowing the benefits of health services, pregnant women run to health facilities for delivery as soon as they face labor without waiting for anyone to take them. [WDA, Seka District]*
>
> … *most of the time, mothers prefer to give birth at home due to a lack of awareness, but those who do have awareness prefer a health center. [MDA, Seka District]*

#### 3.6.2. Knowledge of men identified during labor and delivery

Despite improvements in knowledge about maternal health care, men still have less awareness than women. The improved knowledge of men focused not only on advising and instructing the women to visit health centers but also on escorting her during labor and delivery. The participants have explained the improved knowledge of men as, unlike the previous time, currently men have improved knowledge on the importance of institutional delivery and prefer his wife to give birth in a health facility than at home, unless there is a financial problem.

> *Since the husbands are afraid of problems occurring during pregnancy, they prefer that their wives give birth at a health center. In cases where the pregnant woman needs to give birth at home, the husband takes her to a health center when the labor comes. The husbands never influence or force their wives to give birth at home*. [RL, Gomma District]

> *Husbands are not denying women giving birth at health facilities unless they are poor. [WDA, Gomma District]*

With regard to the low level of knowledge of men about institutional delivery, some men blame themselves after they have observed safe institutional delivery, while his pregnant wife was forced to come to the health facility for delivery services with the support of a neighbor after he refused to let her give birth in the health facility.

> *Even if her husband blamed his neighbors’ for that time—who insisted he carry them to the health facility—he acknowledges them a lot after the pregnant women came to the health facility and attended safe delivery, he said that, “I had lost my child and my wife at that time due to my poor knowledge*.*”*. [HEW, Gomma District]

Another religious leader supported the community’s understanding in general terms by denying the existence of any counter-advocacy for institutional delivery in the community.

> *There are no individuals who are advocating against maternal health services*. [RL, Kersa District]

However, men could prevent pregnant women from attending the women’s network group meeting due to the priority given to his personal interest. The women’s lack of knowledge about their rights and fear of their husbands were associated with these reasons. Despite the women’s netting, it gives pregnant women an opportunity to share their interest and intention about the importance of health facility delivery.

> *“The one thing is that, due to a lack of awareness, they may not have adequate knowledge and do not have education; in addition, they may fear their husband*.*” [Midwifery, Gomma district]*

> *Male farmers want to keep their wives at home to prepare food and warm* ***the house*** *while they return from their work. This can hinder their participation. [PHCUD, Seka Chekorsa District]*

> *There are husbands who say, “****Stay at home, as I [the husband] am coming from my work***.*” We [women’s] are even having such kinds of husbands. [WDA, Kersa district]*

#### 3.6.3. Knowledge of women in post natal care (PNC)

Commonly, due to cultural concerns, most of the women who attended institutional delivery couldn’t come for a postnatal checkup. Besides the culture, the knowledge gap is another contributing factor.

> *All those who gave birth will not come for this service [PNC] on the third and seventh days, according to the new guidelines, even if we encourage them to come*. [PHCUD, Kersa District]

In general, according to the diversified group of participants’ explanation, the level of knowledge of the pregnant women has improved; they now have a clear awareness about maternal health care services and maternal health problems associated with home delivery.

> *“More than anyone, pregnant mothers know the effect of giving birth at home”* [PHCUD, Gomma District]

> *Because they are following different media, they [pregnant women] can tell you many things, because they know the problems they may face*. [HEW: Seka District]

> *“All women in the locality have equal knowledge as health extension workers*.*”* [WDA, Gomma District]

> *“The levels of understanding of the pregnant women have been increasing*.*”* [RL, Kersa District]

#### 3.6.4. Gender-based beliefs identified during delivery and postnatal care services

Gender-based beliefs either facilitate or hinder women’s utilization of maternal health care services and the engagement of men in maternal health care. Culturally, women’s beliefs, particularly during the time of delivery, such as exposing the body of a female to an external person, were considered taboo and prevented them from utilizing services. On the contrary, issues raised were beliefs during delivery associated with: giving birth in a facility to prevent bleeding, saving the lives of the women and the fetus, making the women become clean and netted like brides after delivery, similar to home delivery, and fear of the burden of consequences in the family in case some complication has happened. These were the common beliefs that facilitate maternal health service utilization. Regarding postnatal care, culture-related gender beliefs that couldn’t allow the women to move outside of the home before forty-five days were frequently mentioned as hindering factors for PNC services.

#### 3.6.5. Women’s beliefs about delivery services

There was an indication of a shift in women’s beliefs from giving birth at home to giving birth in health institutions. As mentioned by the participants, institutional delivery alarmingly increased because of a positive transformation of women’s beliefs about maternal health care services.

> *In previous times, many of them gave birth at home. But now, at least more than a hundred women give birth in health facilities every month*. [PUCUD, Seka District]

> *They [pregnant women] are leaving their home for the maternal waiting area before anyone sees them. We [men] simply find them at the [maternal] waiting area. They go to the [maternal] waiting area before getting weak*. [MDA, Gomma District]

Women have a strong belief that home delivery leads to negative health consequences, as does giving birth in a health facility, despite its health benefits. The cleanliness of the care creates an impression, and returning women home like ***“brides”*** helps change their beliefs. Therefore, as participants mentioned, an absolute improvement has been observed in the utilization of delivery services with regards to the belief of being clean, free from bleeding, and free from bad odor.

> *Mostly, they prefer to give birth at a health center, due to the risk of giving birth at home and the safety and care given by professionals at health centers. [MDA, Kersa District]*

> *We [the community] didn’t even know how to stop it well, but they stopped the bleeding properly, and that woman would return home. In this current season, both the bride and the woman who gave birth to the health institutes are similar. [WDA, Gomma District]*

> *Nowadays, women do not compare their shame [due to exposing their private parts] with the benefit of facility delivery, like preventing their lives and their children’s lives from death and any harm. Therefore, they are giving birth at a health facility by considering no loss of blood at delivery and delivering neatly, unlike previous home delivery times, where there was high bleeding up to the extent of spoiling the odor of the women. [HEW, Kersa District]*

According to religious leader explanations, unlike previous experiences, pregnant women in the present prefer to give birth in the health institutes; previously, women who **gave birth at home were an indication of strength**, which used to cure the stigma of weakness given by the societies.

> *At this time, they [the pregnant women] have been using the health center for their own interests. Previously [before the expansion of health facilities], giving birth at a health center was considered shameful. Strong women were the ones who gave birth at home. Therefore, they didn’t give birth at the health center*. ***[RL, Gomma District]***

There was a tendency to be shy about giving birth in the health institute, especially if the health professionals were male, which was associated with a culturally private part where exposure to others was not allowed. But, now things are changed, as long as they understand the benefit of saving the lives of the women and newborns, preventing bleeding and becoming clean and neat after delivery. And therefore, most pregnant women have shown an intense interest in giving birth in health institutions, regardless of the gender of the health professionals.

> *There is improvement [in the health institution] gradually, and now a day’ women are not comparing shame with the benefit of facility delivery, like preventing their and their children’s lives from death and any harm. Therefore, they are giving birth at a health facility by considering no loss of blood at delivery and delivering neatly, unlike previous times where there was high bleeding up to the extent of spoiling the odor of the women*. ***[WDA, Kersa, and Gomma districts]***

#### 3.6.6. Men’s belief identified during delivery service

Some men have the belief that the negative health outcome occurred to the women and the newborns as a result of health facility delivery vis a vis all the previous home deliveries didn’t create any health problems for the women or the newborns.

> *Even if she came and delivered at a health center, if there is a problem with the mother or child, they will probably consider this a mistake done by health professionals or as if they murdered his wife or child. They will try to justify this by stating that she had given these numbers [of children born at home and healthy children] at her home, but now this is the result of her coming to the health center*. [HEW, Gomma District]

Men have shown some sort of reluctant involvement in his wife’s maternal health care services associated with the strong religious belief; they perceived that ***“Allah”*** was protecting from any harmful health consequences and that the women’s and children’s health was protected by the will of ***“Allah”*** rather than modern medicine, therefore they lost interest in pregnant women attending the delivery service. As a PHCUD mentioned, such types of beliefs were a challenge.

> *…Then I asked whether his wife attended delivery service for the current pregnancy, and he [the pregnant woman’s husband] replied she didn’t. When I asked the reason, he related the issue with religion, and he said protection of one’s health was not due to getting modern medication; Allah is the one who protected us from any diseases. …We faced such similar challenges*.*”* [PHCUD, Seka District]

However, the community has an abiding rule, as the participants have mentioned: men have a responsibility to support the women who give birth in the health institute; otherwise, he could be accountable for any undesired health outcomes for the women and the newborn baby if she gives birth at home. This structural protection, besides all the men’s beliefs about maternal health care services, helped him avert his beliefs towards institutional delivery services.

> *He [the husband] will be responsible for any harm that will happen to that [pregnant] woman if she does not use the [maternal] service during pregnancy and delivery*. [Women who gave birth, Kersa Disrict]

> … *The husband will be the one responsible and accountable for her if anything happens to that pregnant woman, including after her death, if it happens*. [Women who gave birth, Gomma District]

> *There are rules and regulations taken as legal ways established at shane and gare [locally assigned names for women association] levels to punish such kinds of crime, and the husband will be asked based on that placed rule if his wife has given birth at home*. [WDA, Seka District]

> … *the husband allows his wife to use health services out of fear of the rules*. [WDA, Kersa District]

> … *Husbands are not denying the pregnant women the right to go to a health facility while they face labor; rather, they take them to the health facility*. [WDA, Seka District]

The men in the current study regret those mothers who have passed away due to a lack of awareness and inaccessibility of maternal health services. Unlike past experiences, currently pregnant women have an opportunity to get adequate health information, either through discussions at home and in health facilities or from different modes of delivery that empower and improve the health literacy. The strong belief that husbands rely on the discussions pregnant women conduct with different parties significantly contributes to save the lives of both the women and the newborn baby.

> *Previously, our [pregnant]mothers due to the lack of awareness and health institutions, we have lost many [pregnant]mothers and sisters. But after this health center is build and started using health extension workers advise the death of mothers were stopped* ***[Husband, Seka district]***

Most men have a strong positive belief that pregnant women should give birth in health institutions. They believe that *“pregnant women should arrive at the health center with the support of people,” which is associated with the belief that “giving birth is just the same as coming back from death*.*”* [RL, Seka District]

*“Yes; we all [male community members] are with her once the labor begins*.*” We go with her until her arrival at the health center. We will attentively stay there to follow her condition, whether she is being referred or not. We never go to our daily work that day. Every member of MDA, the neighbor, and the community at large should attend. We do whatever is needed; we carry if there is no ambulance. [MDA, Gomma District]*

> …*since most husbands are preoccupied with different activities, they send their wives along with somebody else, like their kids. However, if she is getting worse from her pregnancy, he himself accompanies her*. [MDA, Gomma District]

> *There are husbands who never want to separate from their wives, even for one day; who never want to leave them alone. Despite their awareness of the importance of maternity stay; they believe that their wives shouldn’t leave them alone and stay there because they suspect many things, and as a result, they try to avoid using this service*. [Husband, Gomma District]

## 4. Discussion

As part of the safe motherhood initiative project in Africa, the gender-based analysis from the perspective of the stakeholders from the woreda to the federal level, as well as the role of different health development armies and health professionals in MNCH were addressed by the previous researchers **(21)**. The current study explored the perceptions of health professionals who provided health services, the community health development armies and religious leaders, who believed in knowing the social capital factors such as gender-based social norms, power relations, and psychosocial variations that have an influence on the utilization of delivery and post-natal care services. Well explored the gender-based factors enhances to know the mechanisms through which men’s involvement into maternal health care services, furthermore give a complete picture of gender in maternal health care because the diverse dimension analysis provides an eminent input for policymakers and researchers **(2, 4-8, 31, 32)**.

### 4.1. Individual and collective gender norm

Gender roles at the individual and social levels have the potential to shape pregnant women’s maternal health care-seeking behavior. The individual reproductive role that has been experienced by most pregnant women was an early appearance at the health institutes at the onset of labor. The collective role that women in the community have played plenty of activities to facilitate institutional delivery, which could be categorized into two categories: the first was women escorting pregnant women during the delivery time and preparing coffee and meals, which gave psychosocial support, like reducing feelings of seclusion and overcoming the stressful situation connected with labor and birth outcome. There were women who remained at home responsible for providing care for the kids who stayed at home and making the home attractive and conducive during the postnatal period. Such social support gives pregnant women the confidence to give birth in the health institutes because they may not worry about the kids and the household arrangements while she is there. Similarly, the study in Bangladesh supports that the gender role at the household and community level improves the intention of women’s maternal health-seeking behavior **(25)**.

The men’s individual role as husband was informing the community leader, who is responsible for mobilizing others to carry the pregnant woman either in the traditional way or using the ambulance to reach early the health facility, which is associated with the collective male role. Such a role of the husband has a significant contribution to early reaching out to the health institutions to save the life of the woman and the fetus. However, in most of the studies conducted on male involvement during delivery time, accompanying and staying with the women in the health facility frequently described men’s roles, which didn’t indicate the motivation and intention of men about institutional delivery **(19)**. The qualitative study on African American parents in the USA mentioned that there is equal investment and interest in having a child, and that the required responsibility of having the child is willingly shared between the two parents **(26)**.

### 4.2. Gender based power relation

The most commonly observed sensitive activity associated with maternal and child health outcomes was gender-based power relations to determine place of delivery **(25)**. In the current study, shared decision-making was most commonly used to determine the place of delivery, which was often the health institute. In the process of making decisions, sometimes families, neighbors, and health professionals were involved. On the contrary, a qualitative study conducted on different stockholders from the woreda to the federal level in Ethiopia mentioned women have no power in decision-making, even in their reproductive health **(21)**. Despite the fact that the study was part of the IMCHA project, the participants may not have correctly identified the existing power relations the further they are from society. However, the current study’s findings revealed that pregnant women were the last decision-makers, particularly to give birth in a health facility, if they were unable to reach an agreement with their husband. As most of the participants indicated, women have legitimate power, and nothing prevents them from making the free choice to give birth in a health facility. Similarly, in Tanzania, women were the last decision-makers to utilize institutional delivery independent of their husbands’ approval **(33)**. Through time, women’s decision-making capacity has grown, and men have also accepted the right of women to make decisions, particularly on the health condition of themselves and their fetus. The improved systems for delivering maternal health care messages have made an enormous contribution.

This is basically a result of improved knowledge among women through their participation in the health development network, which gives them the confidence to make their own decisions and is mostly supported by their partner on the day of delivery. However, the finding is incongruent with the qualitative study conducted in Ghana, as community opinion leaders mentioned that men have made decisions about the maternal health care service and control all the financial resources. Also supported by health professionals, men give little or no support to pregnant women **(34)**. In India, women make more decisions than men **(35)**. In another study, a man was considered the decision-maker for maternal health care services **(36)**. This may be related to how men’s supremacy in the local area is unbroken. However, many health information dissemination mechanisms exist in the current study area, such as HEW and the Health Development Army (HDA), which play an important role in educating women about their maternal health care rights. That is congruent with the WHO recommendation allowing male involvement in maternal health care, provided they respect, promote, and facilitate women’s choice and decision-making autonomy **(9)**.

In rare situations, the husband dominates the last decision-making, often times associated with advice and encouragement for institutional delivery, although they have the pregnant woman’s acceptance. But the intention of the men was not only to prevent the women from morbidity and mortality but also to protect them from the burden loaded on them in case the women passed away during home delivery. This is supported by the concept of properly understanding the motivational and derived force of men, who play a significant role in addressing gender inequity that can facilitate and improve maternal and child health outcomes **(24)**.

### 4.3. Gender based knowledge variation

The knowledge of both men and women has advanced about the importance of institutional delivery. Therefore, the vast majority of the community preferred their particular associations with fear of bleeding, gaining advice, and being free from spoilage by bleeding. Therefore, the pregnant women reach the maternal waiting area early without telling anyone, sometimes even though their husband has found her in the health institution delivery room. A study in Indonesia found that husband knowledge has a significant association with institutional delivery planning **(27)**. In the current study, some participants mentioned home delivery and men’s not are being escorted as a sign of backwardness. Giving birth in health institutes was considered a sign of backwardness because, besides being an expression of modernization, it was a means to exercise women’s rights and have the potential to make decisions about their own health condition. This is in agreement with the study conducted in Malawi, where male involvement is one of the means to express love

**(20)**. The community has its own social norms that require the male partner to assist the pregnant woman in giving birth in a health facility; otherwise, he is considered irresponsible to his family and must accept responsibility for any unintended maternal and new-born health outcomes. which is similar to the study conducted in India, where men have the responsibility to attend institutional delivery **(20)**. Nepal **(15)** is also included. However, male involvement would be reduced if they were perceived as government rules that also stigmatized women who did not have a husband **(20)**. Another study conducted in Malawi described how, the other way round, the belief of men’s involvement would magnify the weakness of women in the peer group and lead to the loss of independent decision-making power of the pregnant women **(29)**. These may be associated with inadequate social attitudinal change about male involvement that motivate institutional delivery **(14)**. This may be associated with inadequate social attitudes about male involvement that motivate institutional delivery **(14)**. Men have improved knowledge about the health benefits of institutional delivery. They didn’t force the women to give birth at home unless they had financial insecurity. This differs from the study conducted in Afar and Kefa, where men make the majority of decisions regarding maternal health care. However, men have a tendency to prevent pregnant women from attending the women’s association meeting, provided that priority is given to their personal interests and the fear that they could have the possibility to identify their rights. This is in agreement with the study conducted in Ethiopia. Afar and Kefa men did not allow the pregnant women to attend meetings with the health development army due to the fear that they could have known their rights **(16, 37)**.

### 4.4. Gender-based belief system in delivery service

There is an absolute shift in the beliefs of women about the importance of maternal health care services. Most women have understood the discrepancy between receiving health services and not at the time of childbirth. This indicates that any observable evidence has the power to alter the women’s beliefs. Besides, these women have experienced spoiled with blood that creates an unpleasant odor while they gave birth at home, but those who gave birth in the health institutes returned home become pretty, as they have expressed ***“looking like brides*.*” This*** can improve the women’s confidence when they return home, as well as help them not feel ashamed when visitors come to congratulate them due to the absence of any spoiled blood-related odor. The other useful belief was that excessive bleeding might not occur when women give birth in health institutions. Even if it has occurred, health professionals are able to stop it. However, these still contradictory beliefs are observed in some women and have never been changed. It is culturally unacceptable for pregnant women to be seen naked in public, which prevents some women from receiving maternal health care services. Furthermore, some women believe that if their mother was born without complications at home, the same thing can happen to them, which reduces women’s desire to give birth in health facilities.

Regardless of how men hold contradictory beliefs about institutional delivery, the vast majority believe it is a requirement. To be on the safest side, men encourage women to attend institutional delivery because it is associated with the belief that ***“giving birth is like coming from death*.*” Therefore***, they have to attend a health facility. As well as beliefs related to the improved women’s maternal health literacy, this occurred in various ways to enhance their decision to use a health facility during delivery. On the contrary to this, very few men’s beliefs are connected with religion and say ***“everything is found in the hands of Allah*.*”*** *The* health facility doesn’t have any contribution to save the lives of women and newborns. As literature describes, besides the observable experiences of men such as physical support and gender relations, understanding particularly the male partners’ subjective experiences such as belief was not considered as a strategy of men’s involvement in maternal health care **(23)**.

## 5. Recommendation

Support for getting to health centers and managing household activities should be strengthened, so community-created local rules and regulations are critical for improving institutional delivery. The strategy of involving men as a whole is mandatory to avoid delays in the reaching of laboring women to health institutions because it is the husband’s role to inform the community leader early when labor is started. The more husbands understand and believe that giving birth in a health facility improves maternal and newborn health outcomes, the more they will be zealous in informing the community leader as soon as labor begins to obtain support for an early arrival. An improved maternal health service literacy of men and women through sustainable community-based education is critical to enabling them to use maternal health services. Interventions focusing on strengthening the societal values that support utilization of health institutions are crucial, like the household head taking responsibility for any unintended health outcome associated with home delivery and social punishment that shows irresponsibility of the household.

Promoting gender equity at the community level has significantly contributed to ensuring maternal and child health. Therefore, any maternal health strategies and interventions should have to consider the women involved in the local association, which gives an enormous opportunity to discuss any constraints regarding maternal health service utilization. It is obvious that one of the indications of men’s involvement in maternal health is allowing pregnant women to attend such types of meetings. Therefore, it is extremely vital for counseling men to understand that the involvement of women tends to improve as part of the quality of life in the family.

If the research conducted in the future focuses on understanding the mechanisms by which women need support from men as husbands and at the community level, they will properly fill the gap in gender inequity in maternal health care.

## 6. Conclusion

The gender-based roles that the community and health professionals identified have plenty of contributions to promote maternal and newborn health. Therefore, empowered women know their right to maternal health service utilization and have made their own decision. No matter how partners held discussions to identify the place of delivery, the last decision maker, particularly if the pregnant woman intended to give birth in the health facility, As a result, gender-based qualitative data analysis has made a significant contribution to identifying the areas of strength and weakness in maternal health care implementation strategies. Continuous community-level health information dissemination and accessibility of the health facility have the potential to allow the community to produce its own gender-based social norms to promote women’s intentions to seek delivery services. The proper care provided after delivery in the health facility has unprecedented value to the women for repeated use of the service. Because the bleeding that occurred during delivery at home might not be cleaned in a similar fashion to that in a health facility, the odor produced creates discomfort and a loss of confidence in the family.

## Data Availability

All data produced in the present study are available upon reasonable request to the author

## 7. Strength of the study

The current study’s strength comes from including a diverse group of men and women from the community as well as health professionals who have a close relationship with maternal health care. In addition, various data collection methods were used for an in-depth understanding of gender-based maternal health care, particularly during delivery. Moreover, the data was collected from different rural districts, which created an opportunity to triangulate the result and improve the trustworthiness of the finding.

## 8. Limitation of the study

The study has limitations on the detailed exploration of post-natal gender roles, knowledge, and decision-making due to inadequate description by the study participants. The result was based on the participants’ perceived or observed gender-related roles and psychosocial variations, including social support and decision-making in the community, rather than their own. This may cause some social desirability bias.

## 9. Co-authors Contribution

All the co-authors are involved in the LA, AM and MS participated in design of the qualitative study. LA, AM, MS, and YK involved in the process of qualitative guide development for focus group discussion and in-depth interview, as well as the supervision of data collection, manuscript-preparation, and manuscript editing.

## 11. Data availability statement

The datasets that support the findings of this study are in the hands of the authors.

## 13. Ethical clearance

We obtained ethical clearance from the Jimma University College of Health Science Institutional Review Board and the University of Ottawa Health Science and Science Research Ethics Board. All research participants provided verbal consent before they were enrolled in the data collection.

## 14. Acknowledgment

My heartfelt thanks go to Jimma University’s Department of Health, Behavior, and Society, as well as Ottawa University in Canada. My appreciation also goes to all the co-authors and the promoters (Prof. Morankar Sudhaker and Dr. Yohannes Kebede) for their valuable support.

## Appendix

**Table A1.**
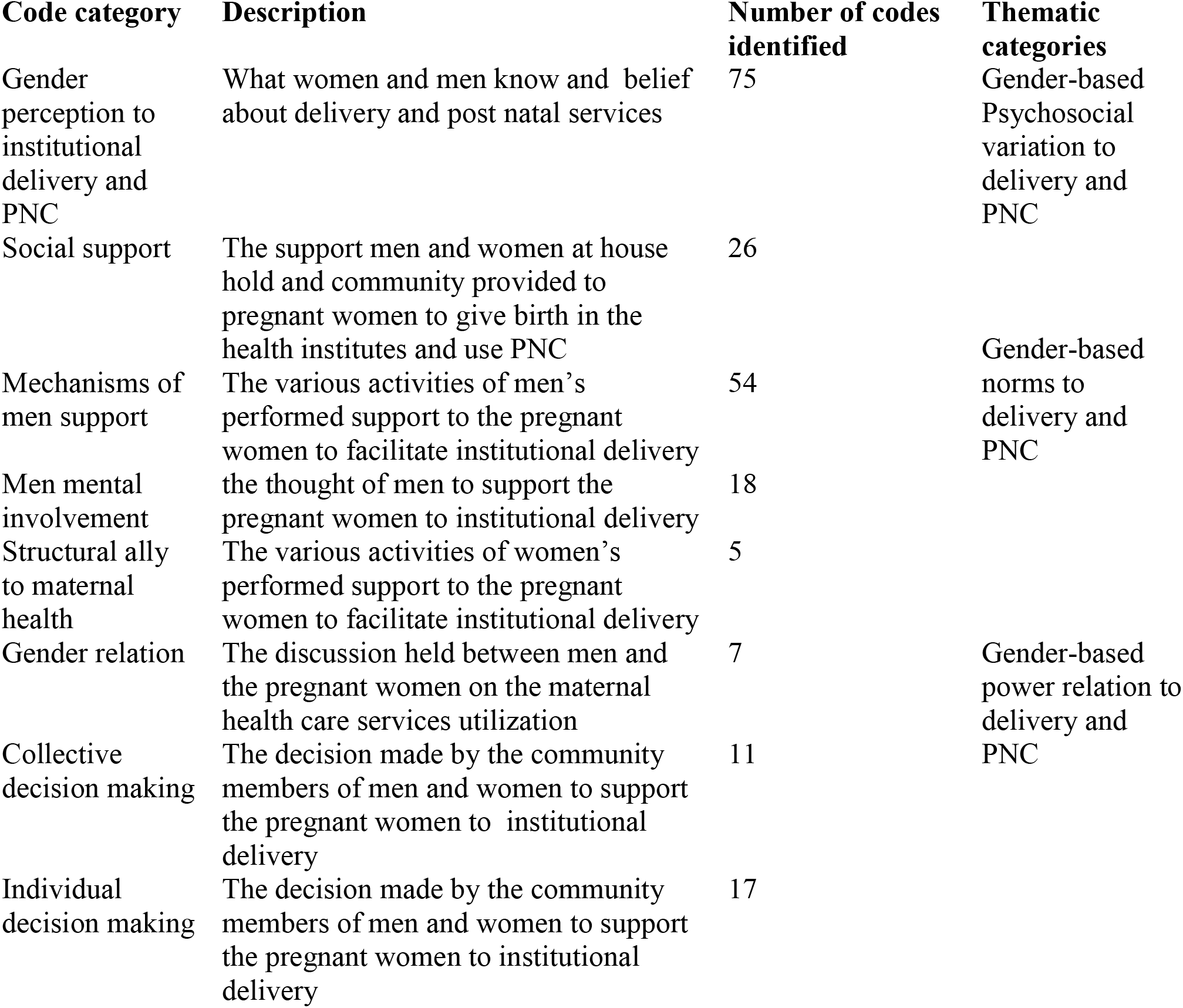
Coding guide applied in the second stage of analysis.

## References

1. World Health Organization. Addressing violence against women and HIV/AIDS: what works?: World Health Organization; 2010.

2. World Health Organization. women and primary health care renewal: a discussion paper: World Health Organization; 2010.

3. World Health Organization. Trends in maternal mortality: 1990-2015: estimates from WHO, UNICEF, UNFPA, World Bank Group and the United Nations Population Division: World Health Organization; 2015.

4. McIntosh CA, Finkle Jljp, review d. The Cairo conference on population and development: A new paradigm? 1995:223–60.

5. Pascoe L, Herstad M, Shand T, van den Heever Ljctsgjn. Building male involvement in SRHR: A basic model for male involvement in sexual and reproductive health and rights. 2012.

6. World Health Organization. Programming for male involvement in reproductive health: report of the meeting of WHO regional advisers in reproductive health, WHO/PAHO, Washington DC, USA 5-7 September 2001. World Health Organization; 2002.

7. Larson ELJEIlLR. United Nations Fourth World Conference on Women: Action for Equality, Development, and Peace (Beijing, China: September 1995). 1996;10:695.

8. Union AJAA. Common African Position (CAP) on the post-2015 development agenda. 2014.

9. World Health Organization. WHO recommendations on health promotion interventions for maternal and newborn health 2015: World Health Organization; 2015.

10. Motala S, Ngandu S, Mti S, Arends F, Winnaar L, Khalema E, et al. Millennium development goals: Country report 2015. 2015.

11. Stipčić A, Ćorić T, Erceg M, Mihanović F, Kolčić I, Polašek OJIjoph. Socioeconomic inequalities show remarkably poor association with health and disease in Southern Croatia. 2015;60(4):417–26.

12. World Health Organization. Health in 2015: from MDGs, millennium development goals to SDGs, sustainable development goals. 2015.

13. Ababa Ajepc. Federal democratic republic of Ethiopia ministry of health. 2003.

14. Barker Gjib. A radical agenda for men’s caregiving. 2014;45(1):85–90.

15. Lewis S, Lee A, Simkhada PJBp, childbirth. The role of husbands in maternal health and safe childbirth in rural Nepal: a qualitative study. 2015;15(1):1–10.

16. Jackson R, Tesfay FH, Gebrehiwot TG, Godefay Hjtm, Health I. Factors that hinder or enable maternal health strategies to reduce delays in rural and pastoralist areas in Ethiopia. 2017;22(2):148–60.

17. Singh K, Bloom S, Brodish PJHcfwi. Gender equality as a means to improve maternal and child health in Africa. 2015;36(1):57–69.

18. Pertierra LR, Hughes KA, Vega GC, Olalla-Tárraga MÁJPo. High resolution spatial mapping of human footprint across Antarctica and its implications for the strategic conservation of avifauna. 2017;12(1):e0168280.

19. Morgan R, Tetui M, Muhumuza Kananura R, Ekirapa-Kiracho E, George Ajhp, Planning. Gender dynamics affecting maternal health and health care access and use in Uganda. 2017;32(Suppl_5):v13–v21.

20. Kululanga LI, Sundby J, Malata A, Chirwa EJAjorh. Male involvement in maternity health care in Malawi. 2012;16(1):145–57.

21. Bergen N, Zhu G, Yedenekal SA, Mamo A, Abebe Gebretsadik L, Morankar S, et al. Promoting equity in maternal, newborn and child health–how does gender factor in? Perceptions of public servants in the Ethiopian health sector. 2020;13(1):1704530.

22. Comrie-Thomson L, Tokhi M, Ampt F, Portela A, Chersich M, Khanna R, et al. Challenging gender inequity through male involvement in maternal and newborn health: critical assessment of an emerging evidence base. 2015;17(Sup2):177–89.

23. Barker G, Das Ajijmh. Men and sexual and reproductive health: the social revolution. 2004;3(3):147–53.

24. Yargawa J, Leonardi-Bee Jjjech. Male involvement and maternal health outcomes: systematic review and meta-analysis. 2015;69(6):604–12.

25. Afsana K, Rashid SF, Chowdhury A, Theobald SJBJoM. Promoting maternal health: gender equity in Bangladesh. 2007;15(11):721-.

26. Alio AP, Lewis CA, Scarborough K, Harris K, Fiscella KJBp, childbirth. A community perspective on the role of fathers during pregnancy: a qualitative study. 2013;13(1):1–11.

27. Kurniati A, Chen C-M, Efendi F, Elizabeth Ku L-J, Berliana SMJHp, planning. Suami SIAGA: male engagement in maternal health in Indonesia. 2017;32(8):1203–11.

28. Gopal P, Fisher D, Seruwagi G, Taddese Hbjrh. Male involvement in reproductive, maternal, newborn, and child health: evaluating gaps between policy and practice in Uganda. 2020;17(1):1–9.

29. Blanc AKJSifp. The effect of power in sexual relationships on sexual and reproductive health: an examination of the evidence. 2001;32(3):189–213.

30. Peneza AK, Maluka SOJGha. ‘Unless you come with your partner you will be sent back home’: strategies used to promote male involvement in antenatal care in Southern Tanzania. 2018;11(1):1449724.

31. World Health Organization. Managing maternal and child health programmes: a practical guide. Manila: WHO Regional Office for the Western Pacific; 1997.

32. Organization WH. Gender, Women and Primary Health Care. A discussion paper 2010.

33. Kohi TW, Mselle LT, Dol J, Aston Mjbhsr. When, where and who? Accessing health facility delivery care from the perspective of women and men in Tanzania: a qualitative study. 2018;18(1):1–9.

34. Craymah JP, Oppong RK, Tuoyire DAJIjorm. Male involvement in maternal health care at Anomabo, central region, Ghana. 2017;2017.

35. Chattopadhyay AJJobs. Men in maternal care: evidence from India. 2012;44(2):129–53.

36. Ladur A. Male involvement in facilitating the uptake of maternal health services by women in Uganda: Bournemouth University; 2021.

37. Kassahun F, Worku C, Nigussie A, Ganfurie GJIJoN Midwifery. Prevalence of male attendance and associated factors at their partners antenatal visits among antenatal care attendees in Bale Zone, South East Ethiopia. 2018;10(9):109–20.

